# THE EFFECTIVENESS OF PRAZIQUANTEL PREVENTIVE CHEMOTHERAPY ON MORBIDITY IN SCHISTOSOMIASIS: A SYSTEMATIC REVIEW AND META-ANALYSIS

**DOI:** 10.1101/2021.11.03.21265867

**Authors:** Reginald Quansah, Mohammad Hassan Murad, Tony Danso-Appiah, Chris Guuri, Alhassan Yakubu, Ann Bretaah Cudjoe, Benson Owusu, Enoch Akyeampong, Nathan Lo, Amadou Garba Djirmay

## Abstract

**Background:** Mass treatment with praziquantel is currently the main strategy used to control the morbidity due to schistosomiasis. Many age groups are at risk of the disease; however, the mass treatment focus mainly on school age children.

**Objectives:** The objective of this review is to estimate the morbidity in at risk groups and identify key groups that should be targeted for preventive chemotherapy treatment with PZQ to control morbidity for schistosomiasis.

**Data sources:** PUBMED and SCOPUS were systematically searched from their inception to 23/11/2020.

**Study eligibility criteria, participants, and intervention:** We searched for studies that evaluated PC in the entire population or a subgroup (PSAC, SAC including adolescents, adults and pregnant women). The Grading of Recommendations, Assessment, Development, and Evaluation approach was used to assess the certainty of the evidence.

**Study appraisal and synthesis methods:** The systematic review followed a pre established protocol. Summary ORs were estimated using the random-effects model. Heterogeneity was assessed by inspecting forest plots and quantified using the I-squared statistic. Subgroup /sensitivity analyses explored heterogeneity.

**Results:** 59 studies met the a priori inclusion criteria and 45 were meta-analyzed. Treatment with PZQ resulted in reductions in right sided hepatomegaly (OR=0.43, 95% CI: 0.22-0.82), hematuria (0.40, 0.25-0.63) hematuria, anaemia (0.70, 0.63-0.79), proteinuria (0.36, 0.22-0.59), and urinary tract pathology (0.37, 0.24-0.56) in SACs; reductions in blood in stool (0.26, 0.14-0.63), splenomegaly (0.56, 0.42-0.55), and urinary bladder lesions (0.26, 0.14-0.49) in entire population were noted. No significant reduction was noted concerning periportal fibrosis, left-sided hepatomegaly, no specific lobe hepatomegaly, and diarrhea prevalence in all age groups. Data were scarce on PSAC.

**Conclusion:** PC with PZQ reduces some infection-related morbidity outcomes in SAC. There is suggestive evidence that PSAC and adult populations may benefit from some outcomes. This assertion was based on low certainty evidence.

**Author summary:** Praziquantel (PZQ) is the main drug used for the mass treatment of morbidity related to schistosomiasis. Current treatment has focused on school-aged children. However, other age groups are also at risk of the disease. We conducted a systematic review and a meta-analysis of 59 studies in PUBMED database (published from inception up to November 2020) to identify key age groups that should be targeted for preventive chemotherapy treatment with PZQ to effectively control diseases related to schistosomiasis infections. Our data suggested that treatment with PZQ in school-aged children led to reductions in certain diseases related to schistosomiasis infection. Pre-school aged children and adults may also benefit from treatment with PZQ. Findings from our study were based on studies with low certainty evidence.

## Introduction

Schistosomiasis is a complex of acute and mainly chronic helminth infections affecting population health and wellbeing. The global burden of schistosomiasis is estimated at 2.5 million disability-adjusted life years (DALYs) [1], and it is transmitted in about 78 countries globally, mostly in developing countries [2]. In 2001, the 54^th^ World Health Assembly (WHA) endorsed resolution WHA54.19 for mass and regular treatment of high-risk groups, particularly school-age children (SAC) as the best strategy, Preventive Chemotherapy (PC) of reducing mortality and morbidity and ensured access to single-dose drugs against schistosomiasis to achieve a minimum target of regular administration of chemotherapy to at least 75%, and up to 100%, of all SAC at risk of morbidity by 2010 [3]. The strategy has a goal to control morbidity through large-scale mass chemotherapy campaigns with PZQ using thresholds of prevalence to categorize at-risk populations and define the frequency of the intervention. The strategy was revised in 2006 to include adults at risk from special groups to entire communities living in high endemic areas [4]. Targets were not met and in 2010 only 30 countries out of 52 requiring PC had implemented mass treatment campaigns with coverage ranging from 4% to 27.5% [5]. The current, revised, guideline endorses regular mass drug administration (MDA) of targeted groups over a few years, to reduce and prevent morbidity and the insurgence of irreversible pathology in adulthood. The guidelines have already been revised a few times in the initiation of the PC strategy 2006 and 2013 [4, 6].

As new evidence emerges, experts have called WHO to develop new evidence-based guidelines and procedures for countries to evaluate the PC strategy. To support the development of these guidelines, we sought to determine the effectiveness of PC in different subgroups of the population and identify the subgroups that would achieve maximum benefit from PC and should be targeted to control severe and subtle morbidity.

## Methods

### Protocol

This systematic review was carried out per methods outlined in the Cochrane Handbook (http://handbook-5-1.cochrane.org). The study questions, the search strategy, and the eligibility criteria were guided by the population, intervention, comparison, and outcome (PICO) statement (http://handbook-5-1.cochrane.org) were developed in consultation with the WHO guideline development group on schistosomiasis. Findings were reported following recommendations of the Preferred Reporting Items for Systematic Reviews and Meta-Analyses statement (PRISMA) (**S1 checklist**) [7]. The Grading of Recommendations, Assessment, Development, and Evaluation (GRADE) approach was used to assess the certainty of evidence (http://www.gradeworkinggroup.org/). The protocol for this review was submitted to the WHO consortium on schistosomiasis for approval. This systematic review seeks to address the question of what population group (s) (Population) should be targeted for the treatment of preventive chemotherapy with praziquantel (intervention) to achieve maximum benefit for the control of severe and subtle morbidity for schistosomiasis (Outcome).

#### Search strategy and selection criteria

We systematically searched for studies in PUBMED (http://www.ncbi.nlm.nih.gov/pubmed) and SCOPUS up to July 2018 using search terms outlined in **S2 Text**. The search was updated in November 2020. We also searched lists of references of systematic reviews [8, 9, 10, 11, 12] and those of relevant studies. **Figure 1** outlines the stepwise approach used in selecting included articles.

**Figure 1.**
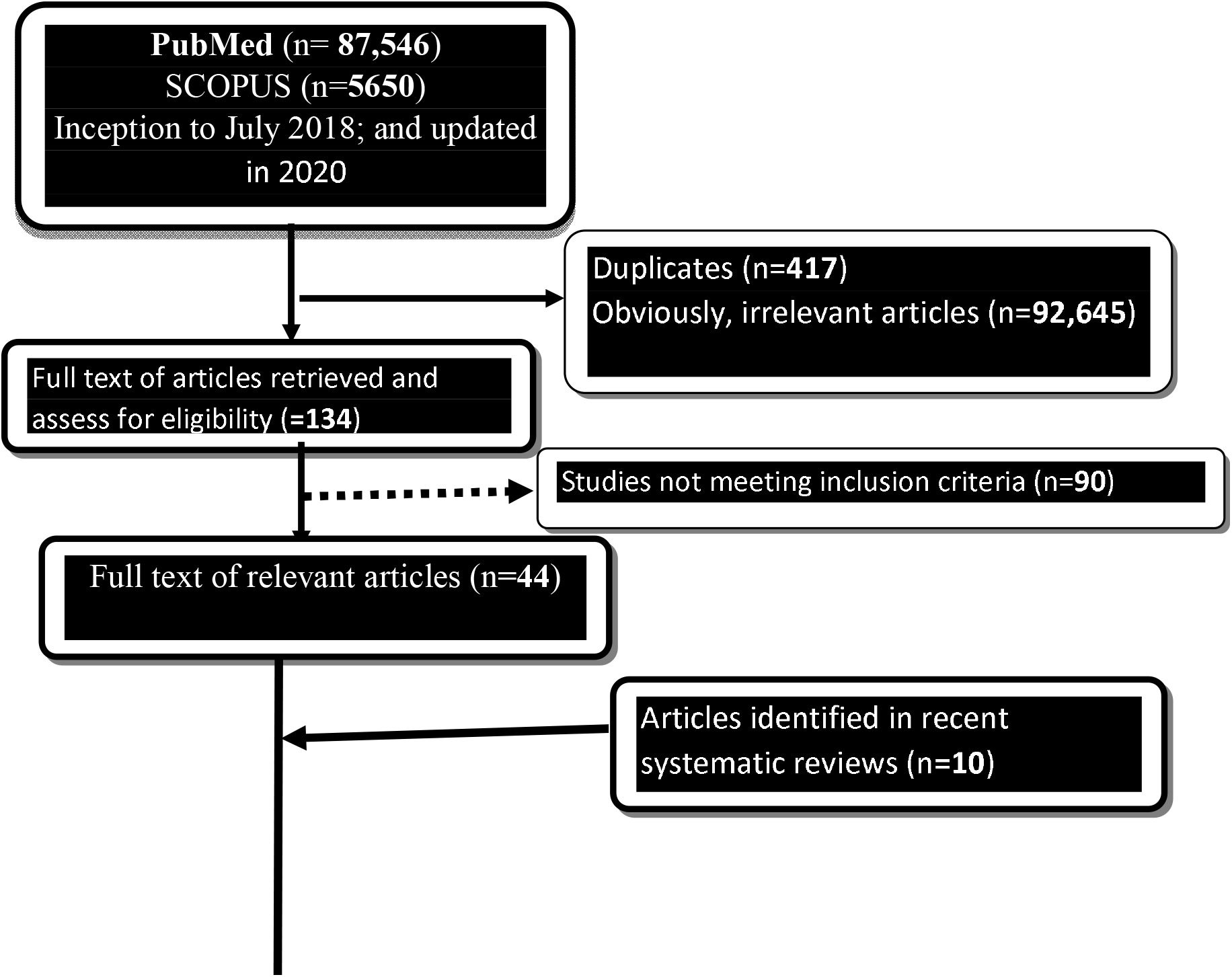
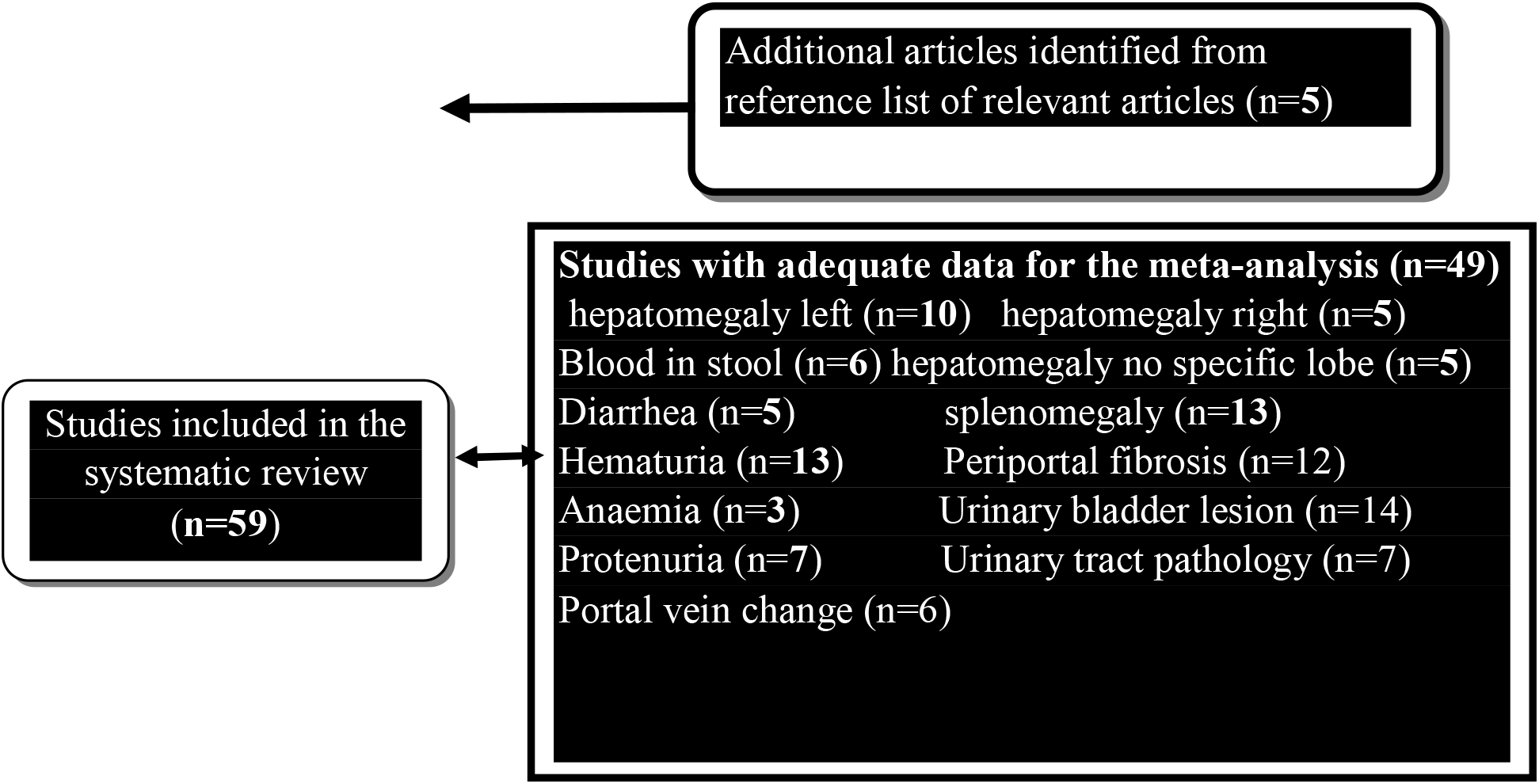
PRISMA study flow diagram showing study selection process.

We included studies (comparative and non-comparative) among the entire community or subgroup (adults including pregnant and lactating women) or high-risk groups or children (e.g, PSAC, SAC, children including adolescents) living in schistosomiasis endemic settings with or without infections of any schistosome type and treated with praziquantel of any formulation and brand at a single/repeated oral dose of 40, 50, 60, 70 and 80 mg/kg alone or in combination with drugs such as those used for the treatment of lymphatic filariasis or malaria. The following outcomes were considered: advanced (e.g., hepatomegaly, splenomegaly, portal fibrosis, cirrhosis of the liver, colonic polyps, abnormalities in the urinary bladder, and lesions of the upper urinary tract) measured by ultrasound, or by standard international classification or other standardized methods [13], or early (e.g., bloody diarrhoea, blood in urine, painful urination, proteinuria, and anaemia) stage morbidity, or subtle morbidity (e.g. cognitive ability, psychometric tests performance, school attendance, and achievement and anthropometric test results) [14]. To provide a comprehensive review of the evidence on the subject, we did restrict intervention studies by type of comparator used. We identified all relevant studies on human subjects with no restriction on language. We excluded reviews, case reports, case series, or animal studies. Intervention studies controlling schistosomiasis infection through access to safe drinking water, provision of improved sanitation facilities, promotion of good hygiene practices, and snail control were excluded.

#### Study Selection and data collection

Titles and abstracts retrieved in the search strategy were reviewed by (RQ). The full text of the remaining articles was retrieved. Using our a priori eligibility criteria four Ph.D. students who have received training in systematic review methods independently identified each potentially relevant studies. A pre-tested data extraction form was used to extract data from relevant studies independently. They also assess the risk bias and evaluate the certainty of evidence with GRADE. Any disagreement was resolved among them, and if not, by (RQ). The following data were extracted from the articles: first author, title, publication date, journal, year of publication, volume, issue, page number; study population characteristics and type, study location and setting, inclusion and exclusion criteria, the total number of participants, recruitment procedure, total number recruited for the intervention arm, total number recruited for the control group, other relevant sociodemographic information, description of the intervention such as dosage, route of administration, type of control, whether PZQ was given alone or in combination, outcomes assessed, diagnostic criteria, and study design.

Extracted data from the articles were entered into an Excel spreadsheet. Each study was assigned a unique Endnote number. These numbers are included in tables and figures, where appropriate, to aid study identification.

### Data synthesis

Effect estimates extracted from eligible studies were odds ratios (ORs) and their 95% CIs. When data were insufficient for meta-analysis, the evidence was summarized narratively. The random effects model was used for meta-analysis because of anticipated heterogeneity in study settings and population [15]. Heterogeneity was assessed using the Q- (p < 0.1 considered significant), and I^2^-statistic (where I^2^-statistic > 50% indicates high, 25–50% moderate, and < 25% low heterogeneity). To explore sources of heterogeneity, the analysis was stratified on a) population type [i.e. adult vs. PSAC vs. SAC vs. entire population) and intervention type (administration of PZQ alone vs. PZQ in combination with other drugs), schistosome species type, etc. A sensitivity analysis was carried out restricting the analysis to studies adjudged to have low/moderate risk of bias [16]. Where data permit (>4 studies or data) publication bias from funnel plots and trim-and-fill methods were applied. The potential impact of missing studies in the funnel plot was assessed with the trim-and-fill methods. STATA software version 15 (StataCorp, College Station, TX, USA) was used for the analysis.

### Assessment of risk of bias

Cochrane Risk of Bias tool (for RCT/CCT) [17] and ROBINS-I (RoB-I) (for non-RCTs) were used to the assessed risk of bias independently by four Ph.D. students. The Cochrane risk of bias (RoB2) looks at random sequence generation (selection bias), allocation concealment (selection bias), blinding of outcome assessors (detection bias), incomplete outcome data (attrition bias), selective reporting (reporting bias), and other bias (baseline imbalance between groups).

The RoB-I guides users through seven domains of bias in non-randomized controlled studies. Pre-intervention parameters assessed any bias due to confounding or arising from the selection of the study population. For the intervention-related domain, any bias originating from the classification of interventions was assessed. Finally, post-intervention domain assessment considered any deviations from the proposed intervention, missing information, bias in outcome assessment, and biased or selective outcome reporting. For each domain, judgment was made of ‘low risk of bias, ‘moderate risk of bias, ‘serious risk’ of bias, or ‘critical risk’ of bias or ‘no information. A study providing inadequate information for any domain or judged ‘no information’ for any domain is judged ‘serious’ in that domain. A study is judged overall low RoB when it scored low for all domains. A study scoring overall moderate RoB, suggests that the study had scored low or moderate for all domains and at moderate in the least one domain. A study scoring overall serious RoB, suggests that the study is scored at least a serious RoB in at least one domain and no critical RoB in any domain. Overall critical RoB suggests that the score is critical in at least one domain. Authors were contacted for additional information, where necessary. Any differences or discrepancies were resolved between assessors and RQ. No quantitative measures were used to express risk of bias [16].

### Grading the certainty of the evidence

The GRADE approach (https://www.gradeworkinggroup.org/) was used to grade the certainty of the evidence across 5 domains: 1) study limitations, 2) inconsistency of results, 3) indirectness of evidence, 4) imprecision and 5) publication bias. Other factors that were considered include 1) the large magnitude of effect and 2) whether plausible confounding would strength an association. (http://www.gradeworkinggroup.org/). Limitation in studies, incorporated assessment of the risk of bias, with the certainty of evidence downgraded if a meta-analysis of studies of low risk of bias only differed from a meta-analysis of all studies; the inconsistency domain addressed heterogeneity using an 80% prediction interval (overlap 1) to indicate considerable heterogeneity and therefore downgrade the certainty of the body of evidence; indirectness related to how well the PICO in the studies in the meta-analysis reflects the original PICO; imprecision was determined if the confidence interval of the pooled estimate included appreciable benefits and harms or if the sample size was small; funnel plot and Eggers test used to assess funnel plot asymmetry, evidence downgraded only if there was a clear indication of publication bias/asymmetry.

## Results

### Literature search

The step-by-step search of the literature is shown in Fig. 1. 59 studies met the a priori inclusion criteria and were included in the systematic review (Fig. 1). Of the 59 studies, 40 were identified through our electronic search, 14 from systematic reviews and meta-analyses [9, 10, 11, 12], with a further five retrieved from references of included studies. 90 studies were excluded for various reasons (**S3 Text)**.

#### Characteristics of eligible studies

Characteristics of the eligible studies by schistosome type and year of publication are shown in **Table S1**. The studies were conducted in three continents: south America (Brazil (n=1), Venezuela (n=1), and Cambodia (n=1)), Asia (the Philippines (n=3), and China (n=5)), and Africa (Kenya (n=7), Cote d’Ivoire (n=1), Tanzania (n=6), Uganda (n=4), Senegal (n=2), Nigeria (n=1), Ethiopia (n=2), Niger (n=2), Sudan (n=4), Mali (n=4), Zimbabwe (n=2), Burundi (n=2), Zambia (n=3), Congo (n=2), Madagascar (n=2), Ghana (n-1) and Egypt (n=1)). The design of the studies included 8 RCTs and 51 observational studies (controlled and uncontrolled pretest-and-posttest designs and repeated cross-sectional designs). The studies mostly included settings with *S. mansoni* (n=24) or *S. haematobium* (n=18), with some data for *S. japonicum* (n=8) or S. mekongi (n=1). PC treatment with PZQ varies by dose and ranges from a single oral dose of 40 mg/kg (tablet or syrup or suspension) or 60mg/kg; 40 mg/kg or 30mg/kg administered twice or thrice over intervals of 2-6 weeks.

The outcomes reported were parasitological (n=13) or morbidity (n=46). Of these, the majority were from high prevalence settings (defined as prevalence >50%), while 38 studies were from moderate prevalence settings (defined as prevalence 10-50%), and the remainder were from low prevalence settings (defined as prevalence <10%). Strategies to reduce infection related morbidity outcomes represented in this study were mostly targeted at SAC (n=29) or entire population (n=22) with some studies targeting PSAC (n=4) or adults (n=4). Infection morbidity related outcomes included blood in stool (n=6), diarrhea (n=5), hematuria (n=13), anemia (n=3), proteinuria (n=7), left-sided hepatomegaly left (n=12), right sided hepatomegaly right (n=5), hepatomegaly no specific lobe (n=4), splenomegaly (n=13), periportal fibrosis (n=13), urinary bladder lesions (n=14), urinary tract pathology (n=8) and portal vein change (n=6) (**Table S1)**.

#### Risk of bias

The consensus judgment for each domain and overall judgment for RCT/CCT is shown in **Table S2**. One study was judged as low risk of bias, 5 were judged as a moderate risk of bias and the remaining were judged as high risk of bias. The consensus judgments for each domain of bias and overall RoB assessments for studies included in the systematic review and meta-analysis are also shown in **Table S2**. For studies reporting multiple outcomes, the RoB domains did not differ by individual outcomes, therefore a single set of domain-specific judgments was used. Of the Repeated cross-sectional studies, only 2 were judged overall moderate RoB, 4 were judged overall serious RoB, and the remaining were judged overall critical RoB. Concerning before-and-after study designs, none was judged overall low RoB, but 1 was judged overall moderate RoB, 7 were judged overall serious RoB, and the remaining were judged overall critical RoB.

### Effect of PC on morbidity

#### Blood in stool

In total, 6 studies provided 7 data for the quantitative analysis (Fig 2). Only one study was available for PSAC. One study presented 2 cohort for SAC and no significant reduction in blood in stool was observed. But no study reported on adults. Four studies were available for the entire population and a significant reduction in blood in stool was observed (0.26, 0.14-0.49; I^2^-index=69%, p=0.022). Overall, treatment with PZQ was associated with reduction in blood in stool (0.51, 0.27-0.98; I^2^-index=90.1%, p=0.000, n=6 studies, 7 data) (see **Table S3**) (Fig, 2). All the studies were adjudged moderate/low RoB for non-randomized controlled trials and some concerns for randomized control trials, thus, n0 sensitivity analysis was carried out. The funnel plot showed no evidence of publication bias (**Fig S1**) and this was confirmed in the trim and *fill methods* (see **Table S3**). Certainty of evidence for this outcome for key age groups was very low (**Table 1)**.

**Table 1.**
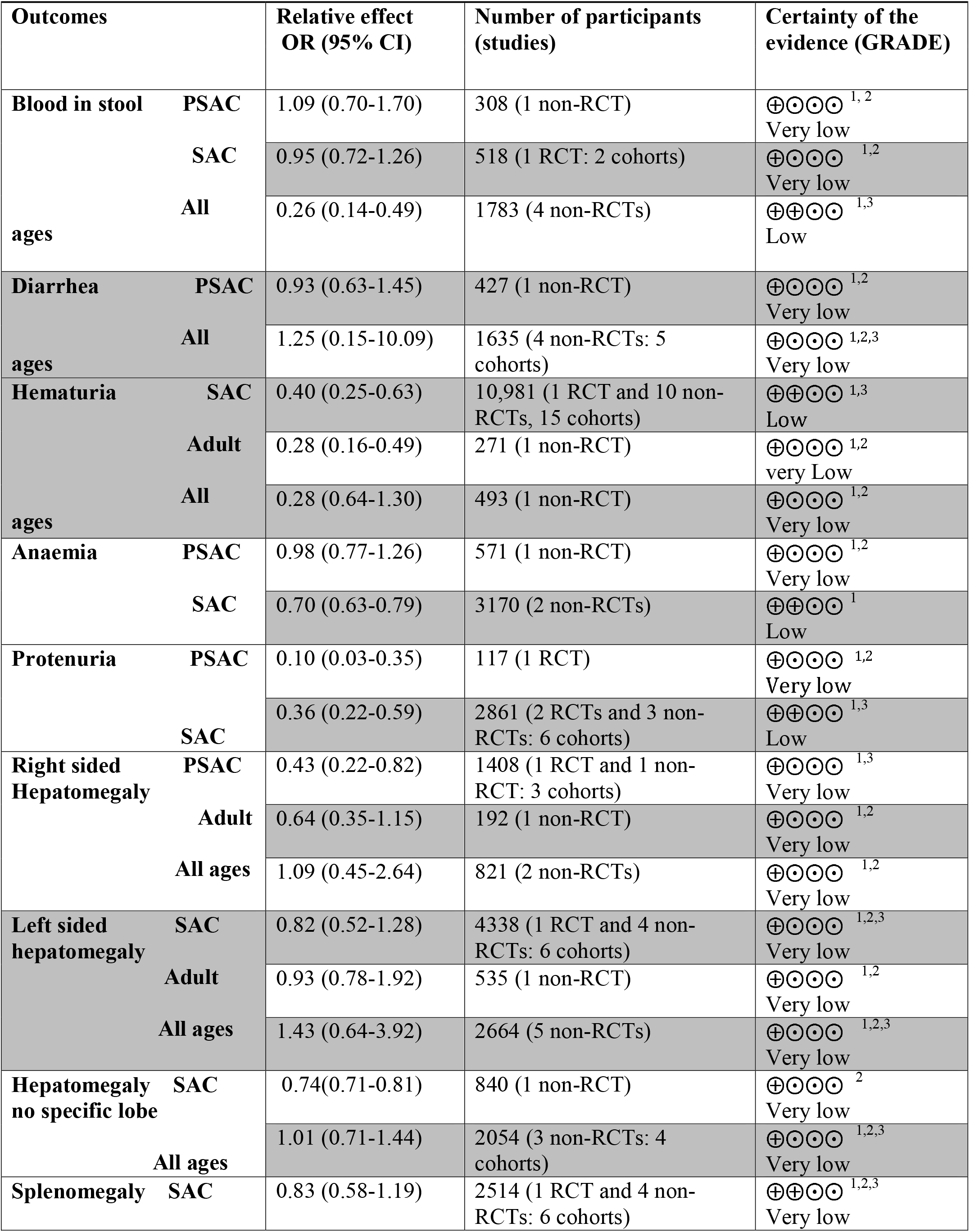
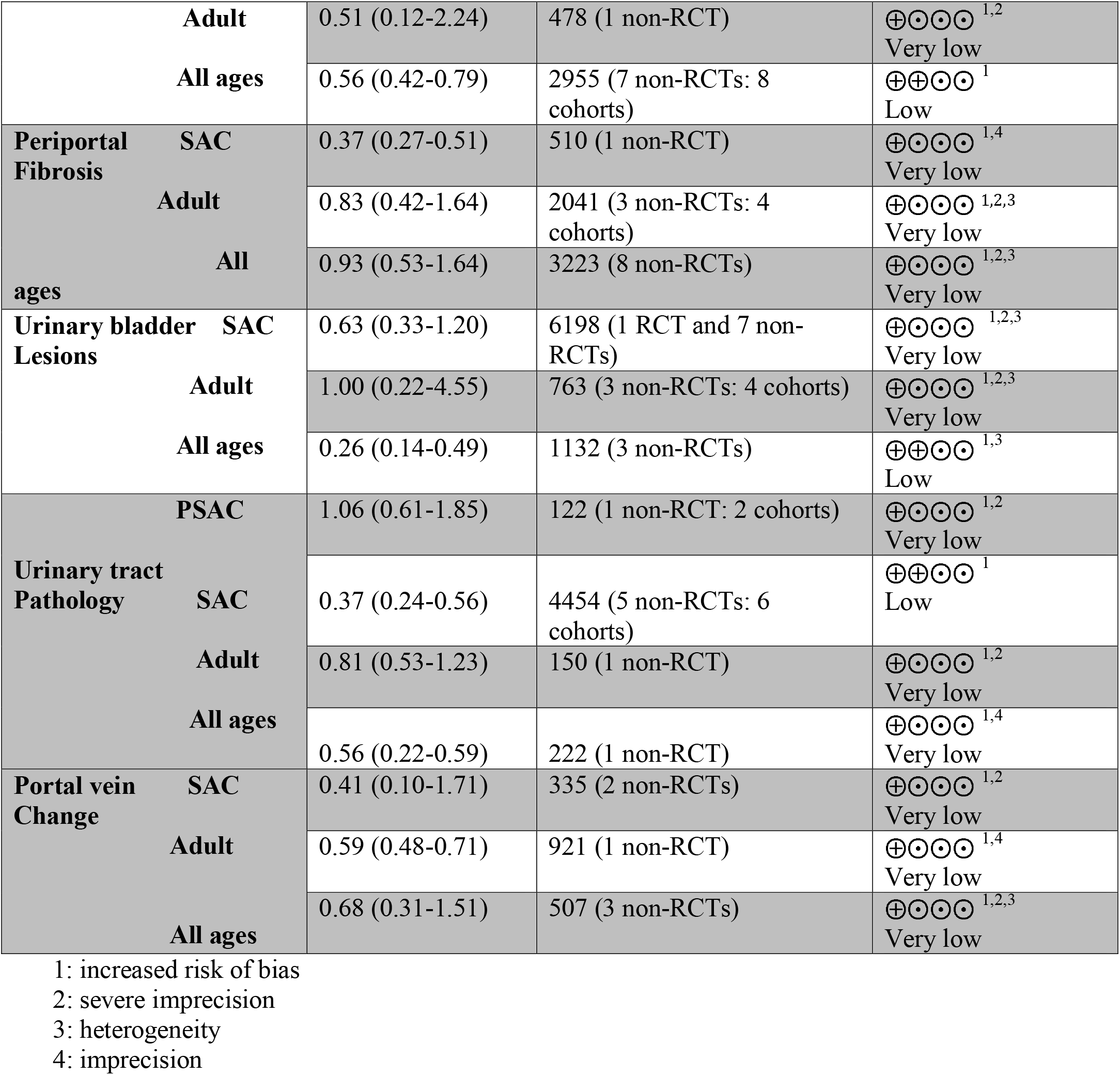
Grading of evidence of the PC treatment with PZQ and morbidity Outcomes.

**Fig. 2.**
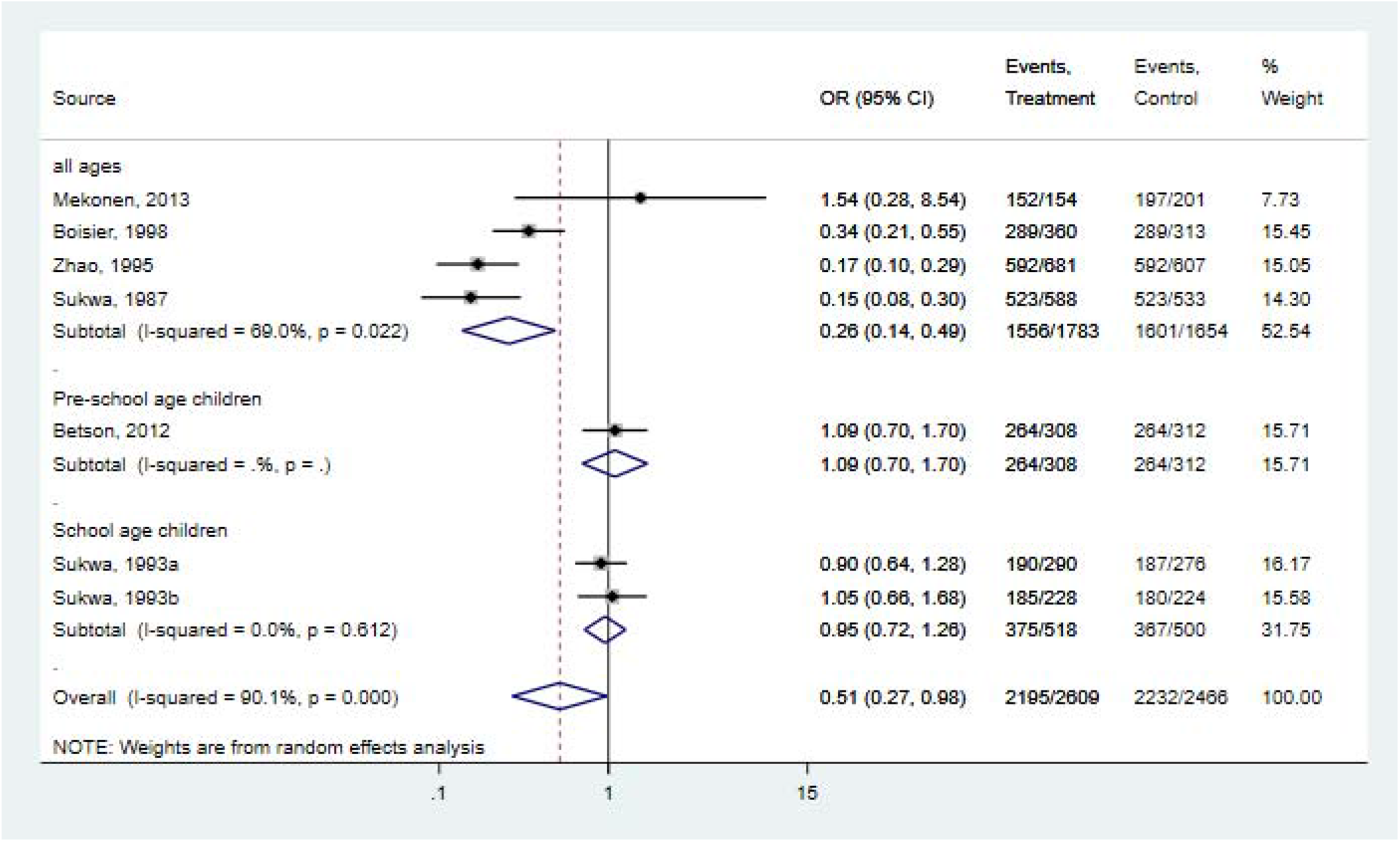
Forest plot for the relation between PZQ/PZQ in combination with other drugs and the risk of blood in stool assessed by population type

#### Diarrhea

Five studies provided 6 data for the meta-analysis on the effect of infection-related diarrhea morbidity (Fig 3). Reduction in diarrhea prevalence was not observed in any of the key age groups including PSAC and the entire population. No study was available on SAC and adults population. The overall analysis did not show any significant reduction in diarrhea morbidity (1.19, 0.22-6.63; I^2^-index=98.5%, p=0.000, n=6 studies) (see **Table S3**). However, PC treatment with PZQ for *S. mansoni* resulted in a significant reduction in infection-related diarrhea morbidity prevalence (0.70, 0.50-0.98; I^2^-index=0.0%, p=0.383, n=3 studies, 5 data) (see **Table S3**). The funnel plot showed no evidence of publication bias (see **Fig S1**) and the *trim* and *fill methods* did not show any material change in the overall effect estimate (see **Table S3**). Certainty of evidence was very low for key age groups (**Table 1**).

**Fig. 3.**
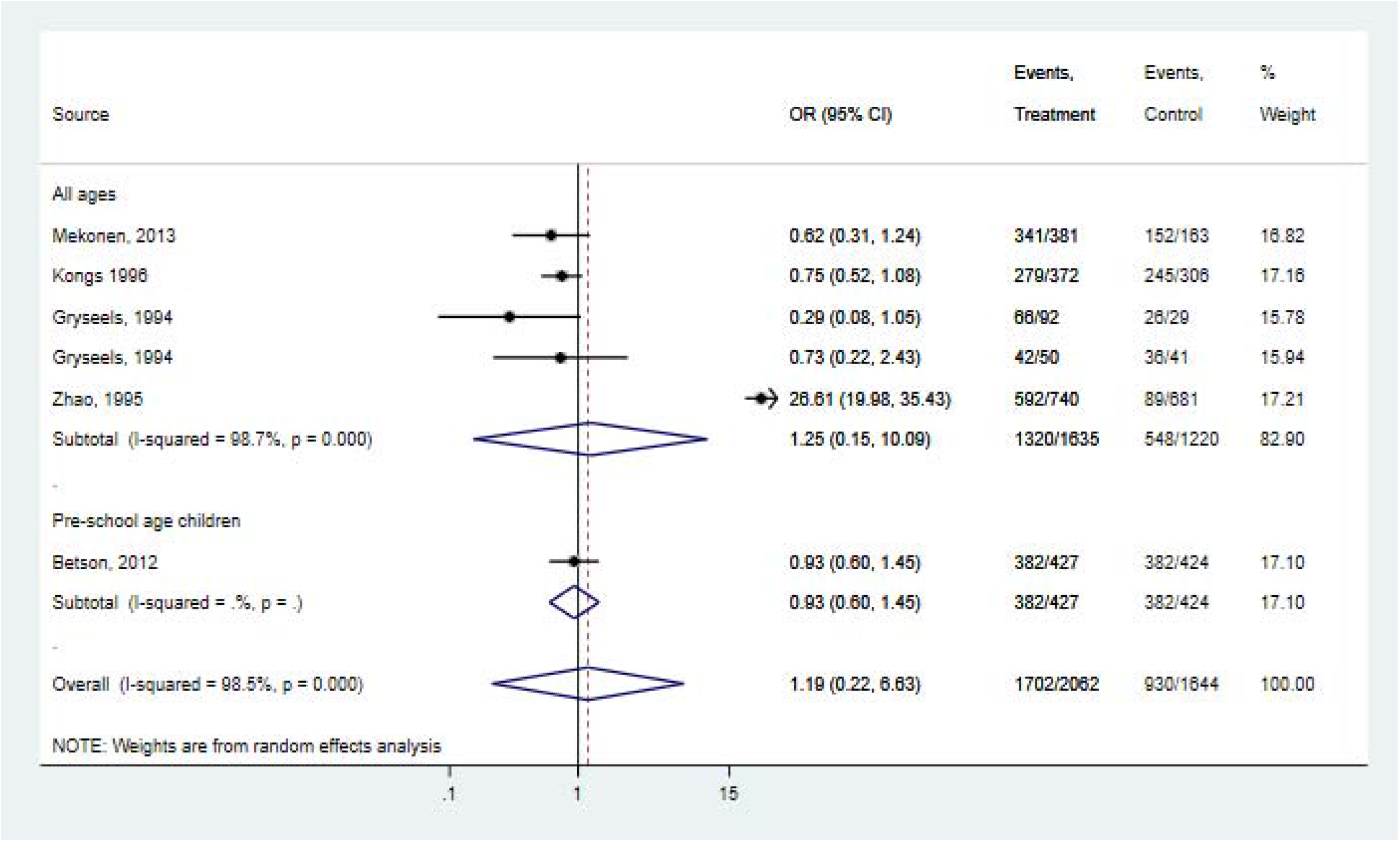
Forest plot for the relation between PZQ/PZQ in combination with other drugs and the risk of diarrhea morbidity assessed by population type

#### Hematuria

Altogether, 13 studies provided 17 data for the quantitative analysis of preventive chemotherapy treatment and infection-related hematuria morbidity (Fig 4). No data was available for PSAC. 11 studies presented 15 data for SAC. There was a 60% (0.40, 0.25-0.63; I^2^index=96.4%, p=0.000, Q-statistics=424.24) reduction in hematuria among this key population. One study each was available for adults and the entire population. Overall, 59% reduction in the prevalence of hematuria following chemotherapy treatment with PZQ was noted (0.41, 0.27-0.63; I^2^index=96.4%, p=0.000, n=13 studies, 17 data) (see **Table S3**). A significant reduction was observed in individuals treated for *S. haematobium* infection (0.50, 0.33-0.74; I^2^index=95%, p=0.000), individuals treated with PZQ alone (0.55, 0.41-0.74; I^2^index=87.9%, p=0.000) or PZQ in combination with other drugs (0.23, 0.08-0.71; I^2^index=98.3%, p=0.000). All the studies were adjudged as critical RoB /high risk of bias on the Cochrane risk of bias tool and thus, no sensitivity analysis was undertaken. The funnel plot did not depict any effect of small studies publication bias (see **Fig S1**) and this was confirmed in the trim and fill method (0.41, 0.27-0.63; I^2^index=96.2%, p=0.000, Q-statistics=422.71) (see **Table S3**). The certainty of evidence was very low (**Table 1**).

**Fig. 4.**
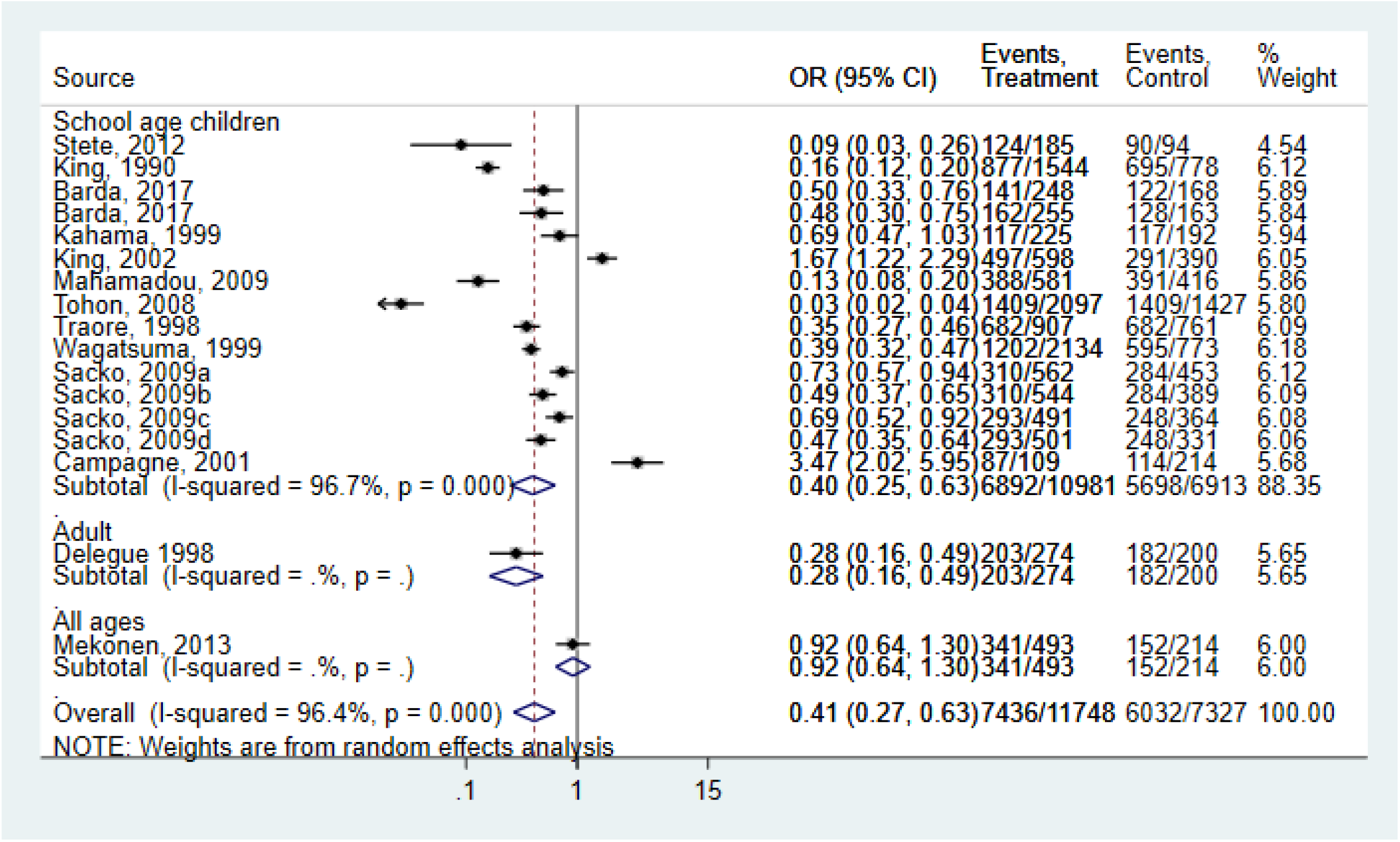
Forest plot for the relation between PZQ/PZQ in combination with other drugs and the risk of hematuria morbidity assessed by population type

#### Proteinuria

Ten cohorts were available from seven studies for the quantitative analysis of the effect of PZQ on proteinuria (Fig 5). One study provided 2 cohort for PSAC, a 90% (0.10, 0.03-0.35; I^2^ -index=76.9%, p=0.000) was noted but the summary OR was unstable. Five studies provided 6 cohort for SAC and reduction in proteinuria prevalence (0.36, 0.22-0.59; I^2^index=80.8%, p=0.000) was observed. No study was available for adults but two studies reporting for the entire population noted a reduction in proteinuria prevalence (0.25, 0.10-0.62; I^2^index=83.4%, p=0.0.014). Again, the summary OR for this population was unstable. Overall, treatment with PZQ was associated with a reduction in proteinuria prevalence (0.30, 0.21-0.44; I^2^ -index=76.9%, p=0.000) (Fig). Administering PZQ alone (0.22, 0.12-0.41; I^2^index=80.3%, p=0.000) or PZQ in combination with other drugs (0.43, 0.03-0.35; I^2^index=0%, p=0.758), administering 40mg/kg PZQ (0.34, 0.23-0.49; I^2^index=78.8%, p=0.000) or 60 mg/kg (0.09, 0.02-0.29; I^2^index=0%, p=0.964) and treating individuals infected with *S. haematobium* (0.28, 0.16-0.48; I^2^index=79.5%, p=0.000, Q-statistics=39.024) resulted in significant reductions in proteinuria prevalence (see **Table S6**). The studies were adjudged as critical or high RoB and thus no sensitivity analysis was undertaken. The funnel plot depicted a small study effect (see **Fig S1**) and confirmation was established in the trim and fill method (0.38, 95% CI 0.26–0.55; I^2^index=74.12%, p=0.000) (see **Table S3**). Certainty of evidence was very low (Table 1).

**Fig. 5.**
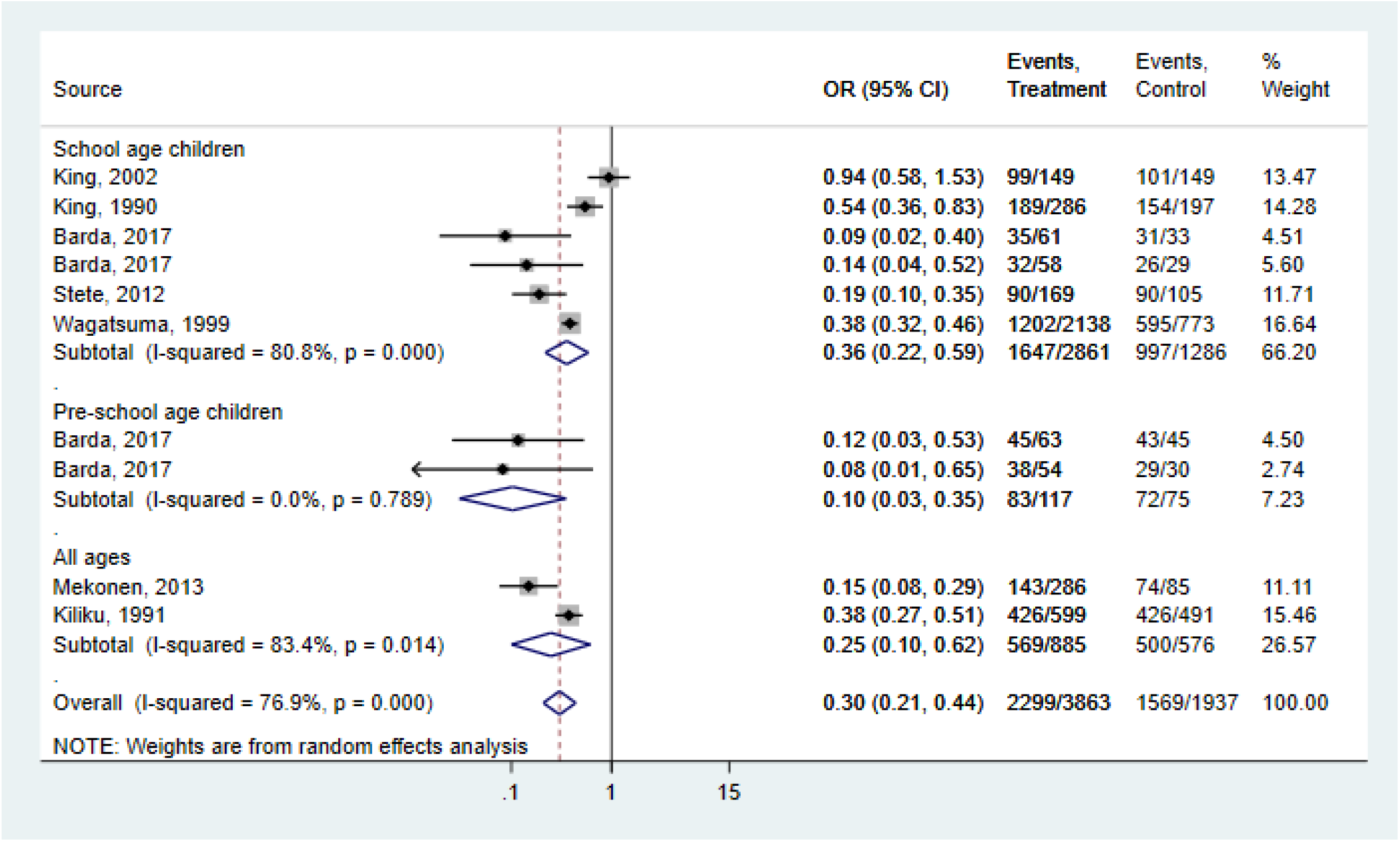
Forest plot for the relation between PZQ/PZQ in combination with other drugs and the risk of proteinuria morbidity assessed by population type

#### Anaemia

The prevalence of anaemia was reported in 3 studies. Only one study reported on PSAC and no study was available among adults and the entire population (Fig 6). Reduced anaemia prevalence was noted in SAC (0.70, 0.80-0.79; I^2^index=0%, p=0.782, n=2). Overall, treatment with preventive chemotherapy resulted in 21% reduction in the prevalence of anaemia (0.79, 0.63-0.98; I^2^index=66.2%, p=0.052, n=3). There was no enough data to explore the source of heterogeneity. Publication bias was also not investigated. Certainty of evidence was very low (**Table 1**).

**Fig. 6.**
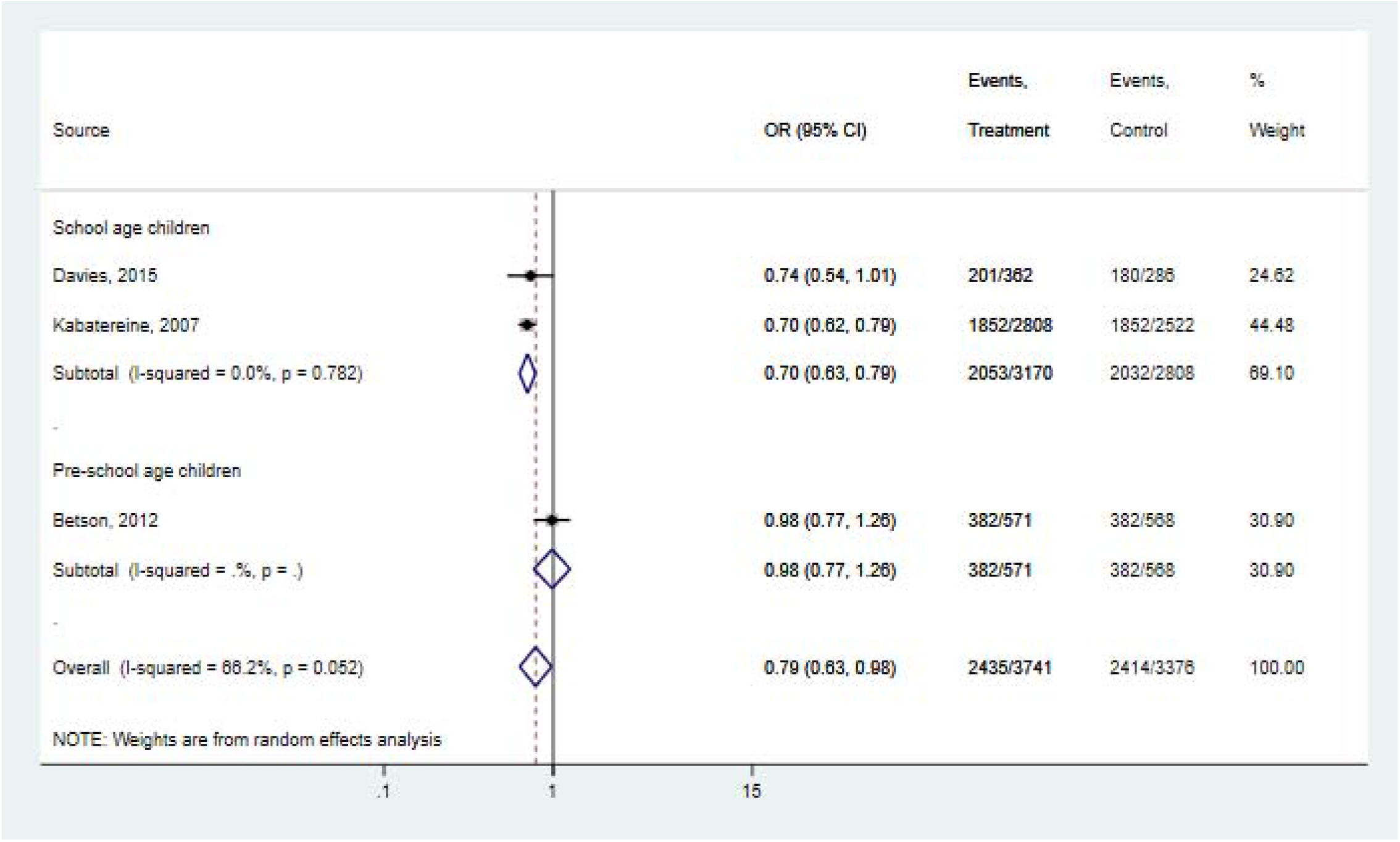
Forest plot for the relation between PZQ/PZQ in combination with other drugs and the risk of anaemia assessed by population type

### Portal vein changes

Six studies provided data for quantitative analysis of the prevalence of the portal vein changes (Fig 7). No data was available for PSAC. Only one study was available for the adult population. Preventive chemotherapy with PZQ among SAC and the entire population did not show any significant portal vein changes. The overall summary OR was 0.59 (0.39-0.87; I^2^index=72.8%, p=0.002, n=6). Preventive chemotherapy treatment with PZQ among SAC and the entire population did not show any significant reduction in the portal vein changes. Treatment of schistosomiasis type and RCT did not show any significant reduction either. But in before-and-after study designs a reduction in infection-related portal vein changes was noted (see **Table S3**). The funnel plot did not show any small study effect (see **Fig S1**) and this was confirmed in the trim and fill method (0.58, 0.26–0.57; I^2^index=78.12%, p=0.000, n=10) (see **Table S3**). Certainty of evidence was very low to low (**Table 1**).

**Fig. 7.**
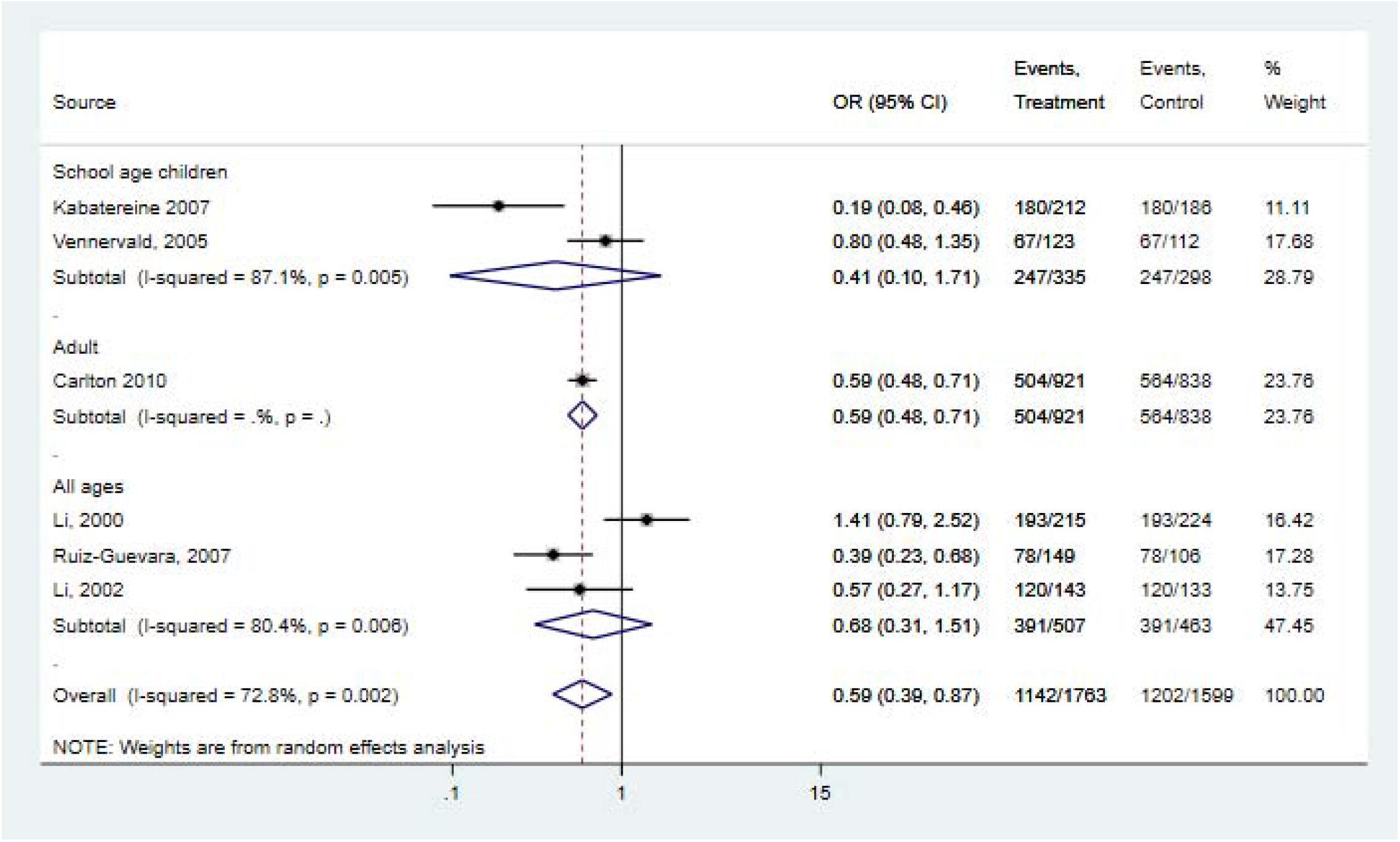
Forest plot for the relation between PZQ/PZQ in combination with other drugs and the risk of portal vein change assessed by population type

**Fig. 8.**
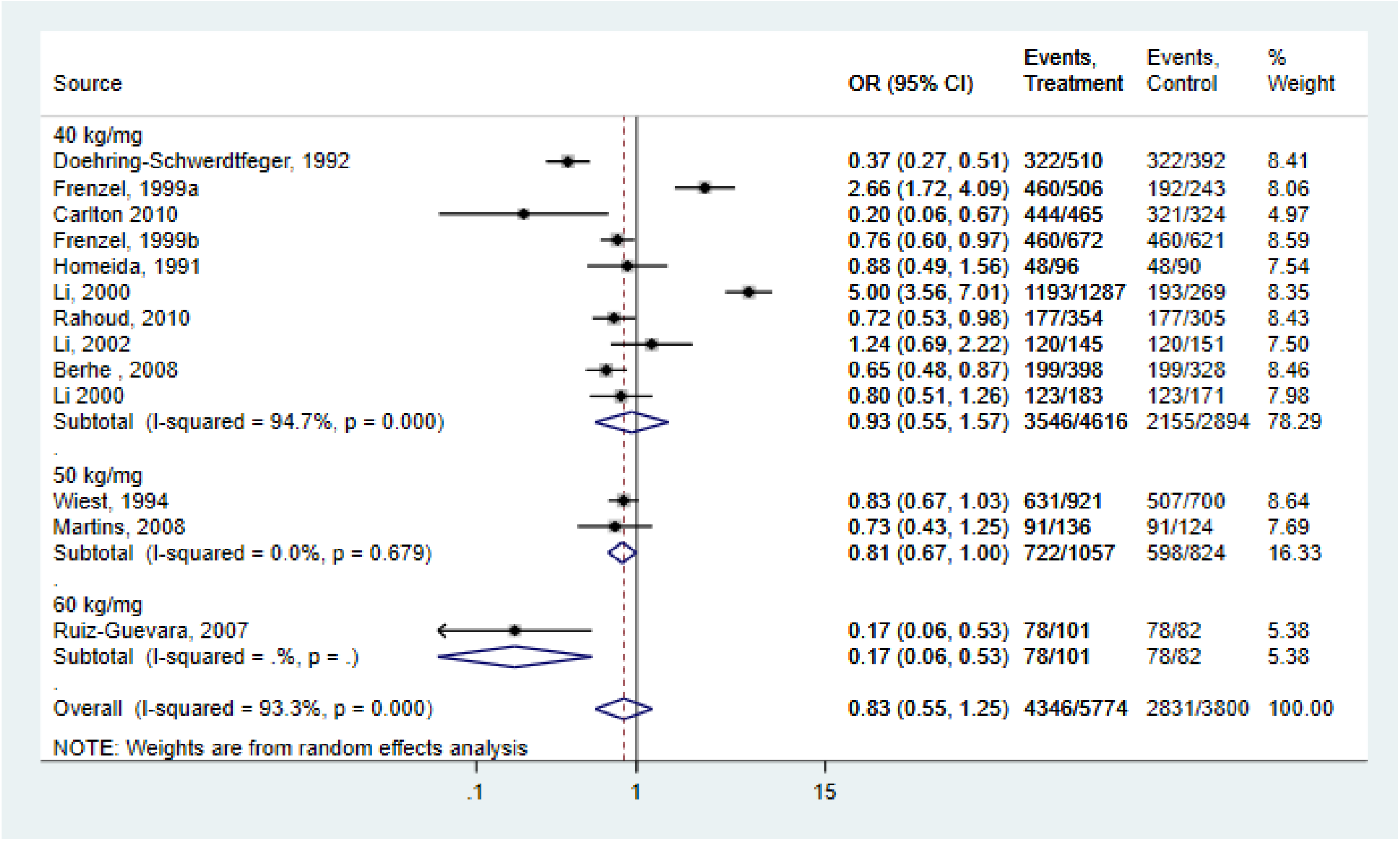
Forest plot for the relation between PZQ/PZQ in combination with other drugs and the risk of Periportal Fibrosis assessed by population type

#### Periportal fibrosis

Twelve studies provided 14 data for the quantitative analysis of periportal fibrosis. Only one study reported on PSAC but no study was available for the entire population. Among SAC (0.93, 0.53-1.64) and adults (0.83, 0.42-1.64) treatment with preventive chemotherapy with PZQ led to reductions in periportal fibrosis but this was not significant. The overall summary OR did not show any significant reduction in periportal fibrosis. Repeated cross-sectional studies reported a 29% reduction (0.71, 0.51-0.99; I^2^index=61,3%, p=0.052, n=4) in periportal fibrosis. one study scored low on RoB2, the remaining were judged critical RoB, and thus, no sensitivity analysis was carried out. A funnel plot shows moderate asymmetry (see **Fig S1**) and confirmatory results were noted in the trim and filled method (see **Table S3**). Addition of three studies did not see any change in the prevalence of periportal fibrosis (0.87, 0.65-1.45; I^2^index=92.7%, p=0.000) (see **Table S3**). Certainty of evidence was explored, and this was very low to low (Table 1).

### Hepatomegaly (without lobe specified)

All four studies provided 5 data for quantitative analysis of the prevalence of hepatomegaly (without lobe specified) (Fig 9). Gryseels et al [18] investigated the effect of repeated preventive chemotherapy treatment with PZQ on *S. mansoni* infection-related morbidity in rural communities in Burundi (Gihungwe and Buhandagaza/Kizina). 40 mg/kg PZQ was administered at 0, 12, 24, and 36 months to individuals displaying eggs in a single 28-mg Kato slide. At each time point, participants were interviewed and subjected to clinical examination and duplicate Kato smear. No study was available on PSAC and adults. Only one study reported on SAC. Among the entire population, the relation between preventive chemotherapy with PZQ and hepatomegaly without lobe specified was not significant.

**Fig. 9.**
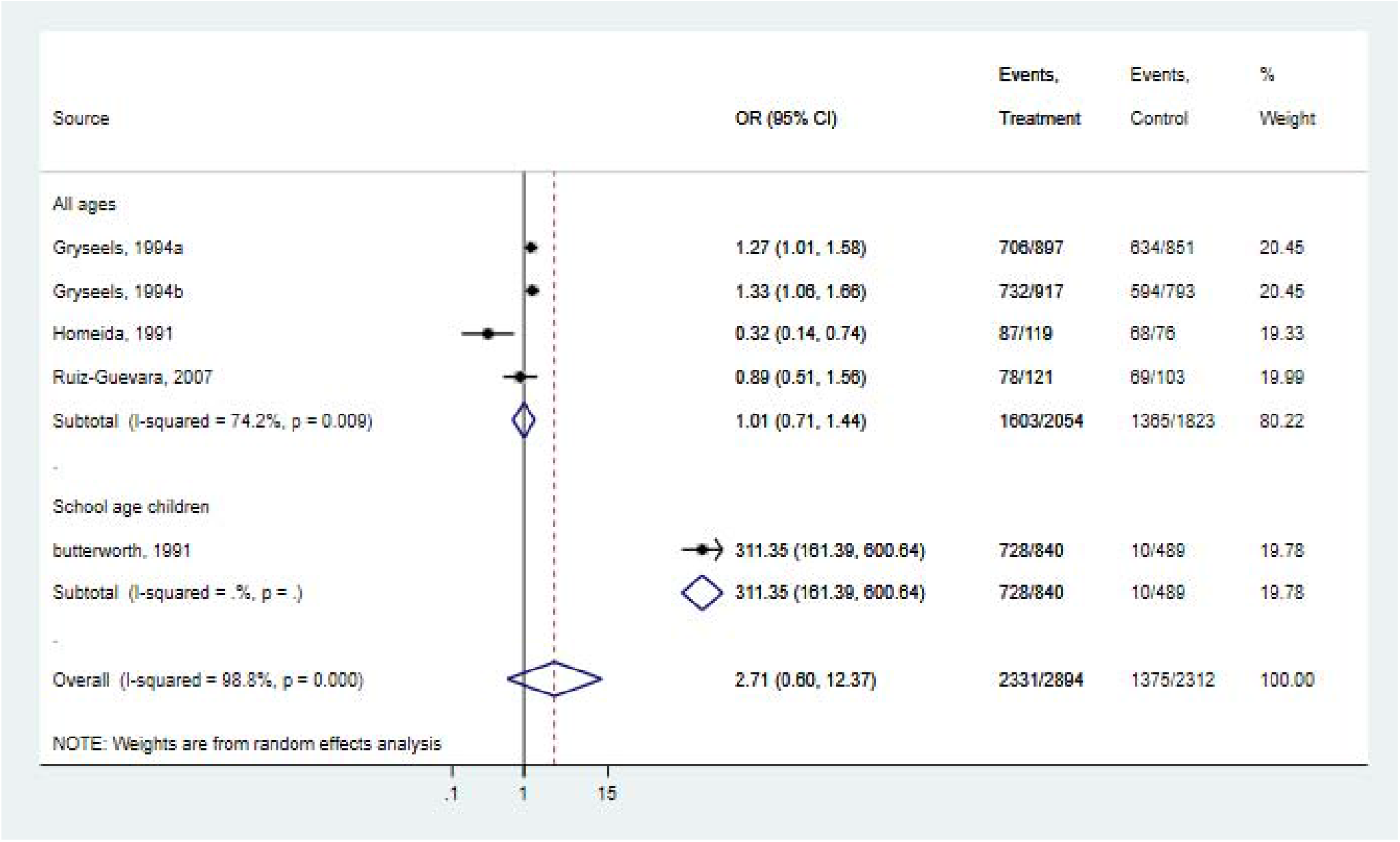
Forest plot for the relation between PZQ/PZQ in combination with other drugs and the risk of hepatomegaly without specified lobe assessed by population type

Overall, preventive chemotherapy treatment with PZQ was not significantly associated with hepatomegaly with no specific lobe. Publication bias was not observed (see **Fig S1**). The addition of two studies in the trim and fill methods did not see any change in the prevalence of left-sided hepatomegaly (see **Table S3**). The certainty of the evidence was very low (Table 1).

#### Hepatomegaly left lobe

A total of 10 studies provided 12 data for quantitative analysis of the prevalence of left-sided hepatomegaly (Fig 10). Only one study was available among adults, no study reported on PSAC. Preventive chemotherapy treatment with PZQ was not significantly associated with hepatomegaly left lobe in SAC and the entire population. The overall summary OR did not show any significant reduction in left-sided hepatomegaly (see **Table S3**). A funnel plot did not indicate any evidence of publication bias (see **Fig S1**). The addition of three studies in the trim and fill methods did not see any material change in the prevalence of left-sided hepatomegaly (see **Table 3**). Certainty of evidence was very low to low (Table 1).

**Fig. 10.**
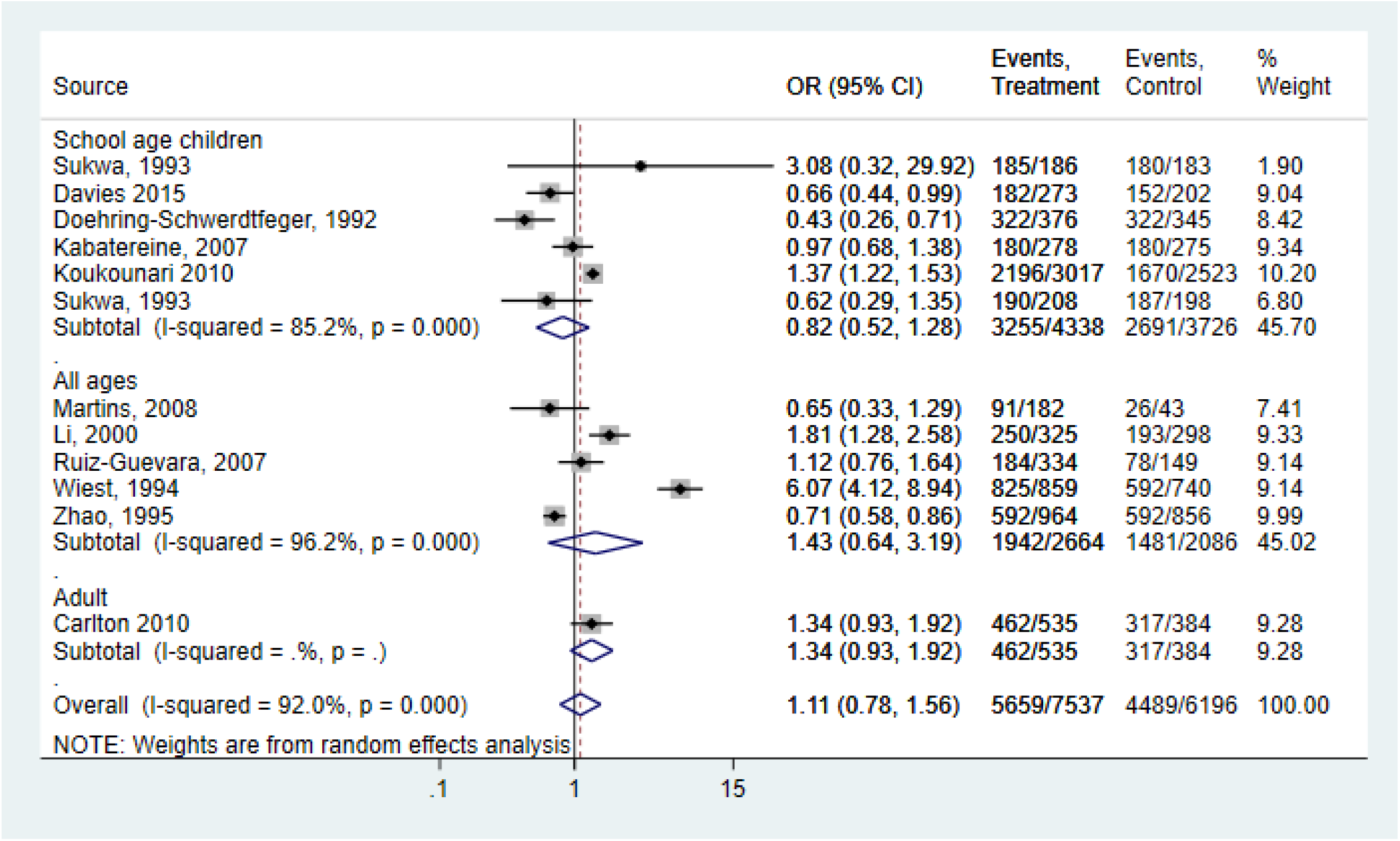
Forest plot for the relation between PZQ/PZQ in combination with other drugs and the risk of hepatomegaly left lobe assessed by population type

#### Hepatomegaly right lobe

A total of 5 studies provided 6 data for quantitative analysis of the prevalence of right-sided hepatomegaly (Fig 11). No study was available among PSAC. Only one study reported on adults. There was a reduction in left-sided hepatomegaly prevalence in SAC (0.43, 0.22-0.82; I^2^index=77.0%, p=0.013, n=4). Preventive chemotherapy treatment with PZQ did not shown any significant reduction in the overall summary OR (0.62, 0.35-1.10; I^2^index=88.3%, p=0.000, n=6). 33% (0.67, 0.47-0.81; I^2^index=0.0%, p=0.915, n=2) and 52% (0.48, 0.30-0.78; I^2^index=70%; p=o,028, n=4) reduction in left-sided hepatomegaly prevalence in patients treated for *S. mansoni* and those treated with PZQ at 40mg/kg respectively (see **Table S3**). Publication bias was not observed. (see **Fig S1**). The was evident in the trim and fill methods (see **Table S3**). The certainty of the evidence was very low to low (Table 1).

**Fig. 11.**
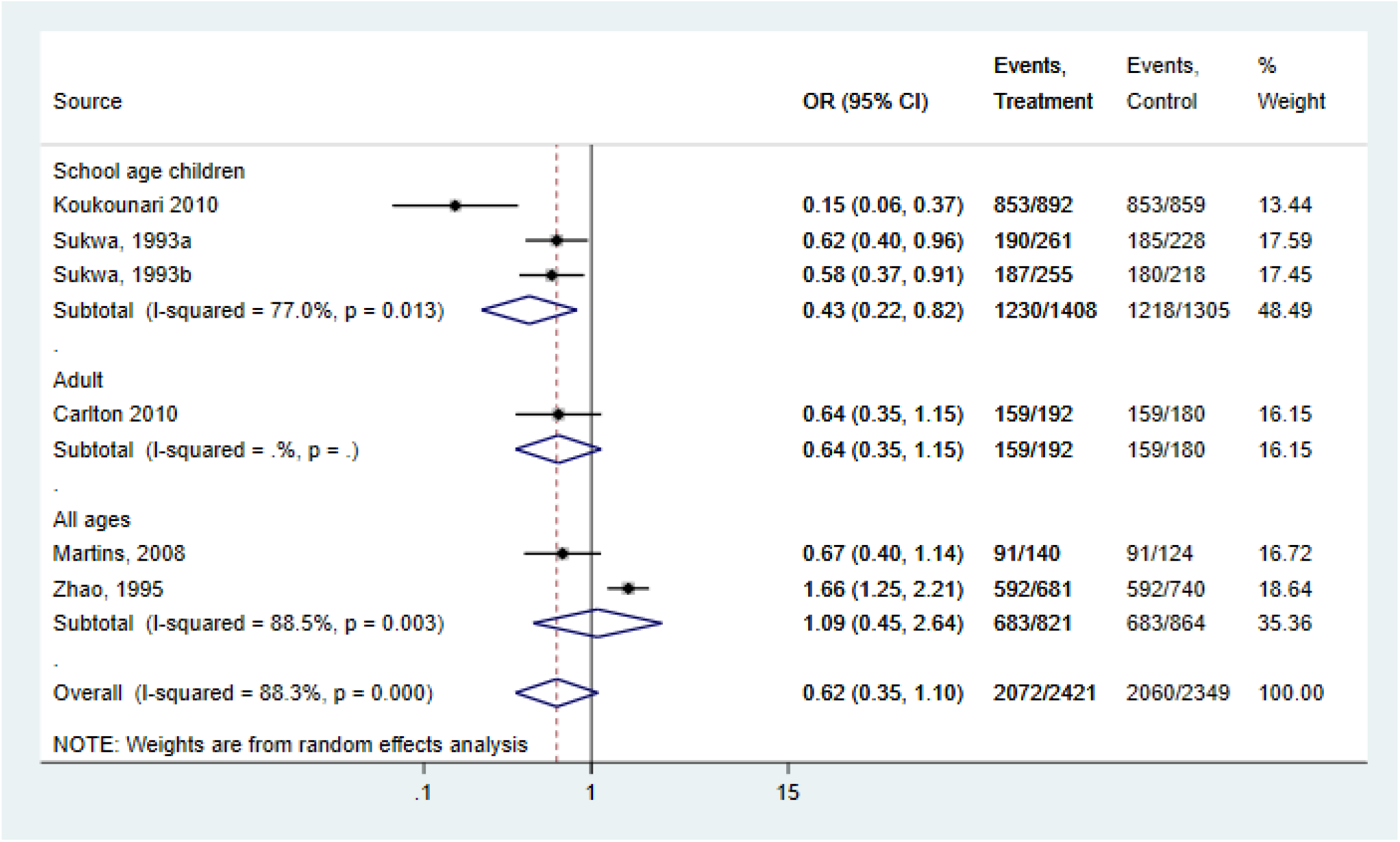
Forest plot for the relation between PZQ/PZQ in combination with other drugs and the risk of hepatomegaly right lobe assessed by population type

#### Splenomegaly

Thirteen studies provided 15 data for the meta-analysis of the prevalence of splenomegaly (Fig 12). Data was no available for PSAC, Only one study reported on adults. Among entire population, 44% (0.56, 0.42-0.75; I^2^index=47.6% p=0.064, n=8) reduction in infection related splenomegaly was observed. No significant reduction was noted in SAC. Overall summary OR showed 33% (0.67, 0.53-0.87; I^2^index=69.9%, p=0.000, n=6) reduction in infection related splenomegaly. This finding was confirmed in before-and-after study designs (0.66, 0.51-0.86; I^2^index=71.8% p=0.000), and individuals treated for *S. mansoni* (0.61, 0.37-0.99; I^2^index=77.7% p=0.000) or *S. japonicum* (0.61, 0.50-0.76; I^2^index=24.2% p=0.266, Q-statistics=3.958) (see **Table S3**). A funnel plot of asymmetry was observed suggesting a potential publication bias (see **Fig S1**) and this was confirmed in the trim and fill method (0.79, 0.61-1.01; I^2^index=73.4%, p=0.000, Q-statistics=63.80) (see **Table S3**). The certainty of evidence was very low to low (Table 1).

**Fig. 12.**
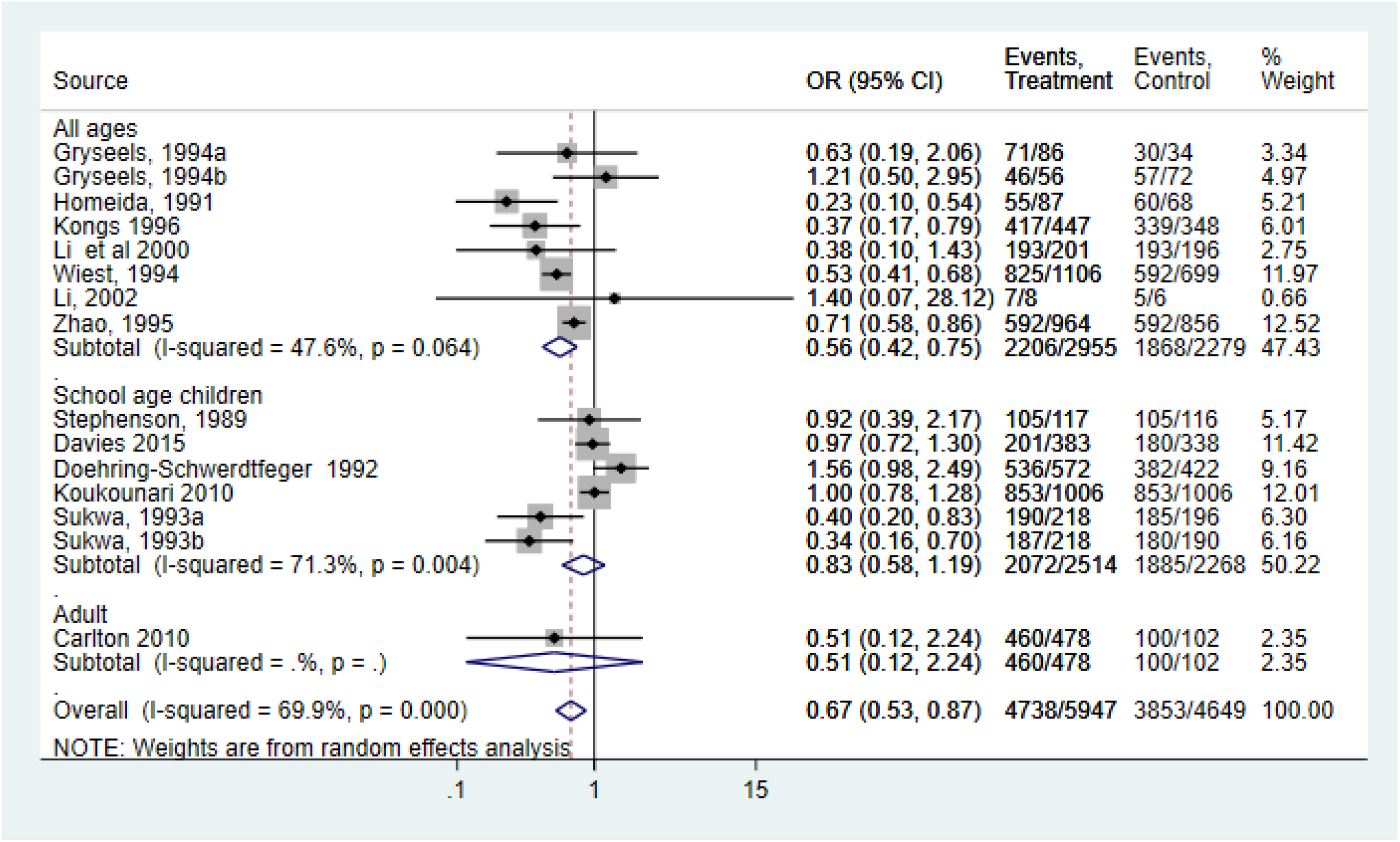
Forest plot for the relation between PZQ/PZQ in combination with other drugs and the risk of splenomegaly assessed by population type

#### Urinary tract pathology

Seven studies provided 10 data for the quantitative analysis (Fig 13). Only one study each reported on the entire population and adults. One study provided 2 data for PSAC and the reduction in urinary tract pathology was not significant. Among SAC, a reduction in urinary tract pathology was noted (0.37, 0.24-0.56; I^2^index=71.9%, p=0.003, n=5 studies provided 6 data). The over summary OR was 0.48 (0.34-0.69; I2index=79.2%, p=0.000, n=7 studies providing 10 data). Before-and-after designs (0.38, 0.23-0.63; I^2^index=84.3%, p=0.000), studies following participants for 11 to 20 months (0.30, 0.19-0.48; I^2^index=76.5%, p=0.005), individuals treated for *S. haematobium* (0.51, 0.32-0.80; I^2^index=78.5%, p=0.000), treatment with PZQ alone (0.47, 0.29-0.78; I^2^index=77.1%, p=0.000) and administration of 40mg/kg (0.44, 0.30-0.66; I^2^index=81.7%, p=0.000) confirmed this finding. Asymmetry was obvious in a funnel plot (see **Fig S1**) and this was confirmed by trim and fill method. Inclusion of 2 studies did not materially changed the overall summary OR (0.48, 0.34-0.69; I^2^index=79%, p=0.000) (see **Table S3**). The certainty of evidence was very low to low (Table 1).

**Fig. 13.**
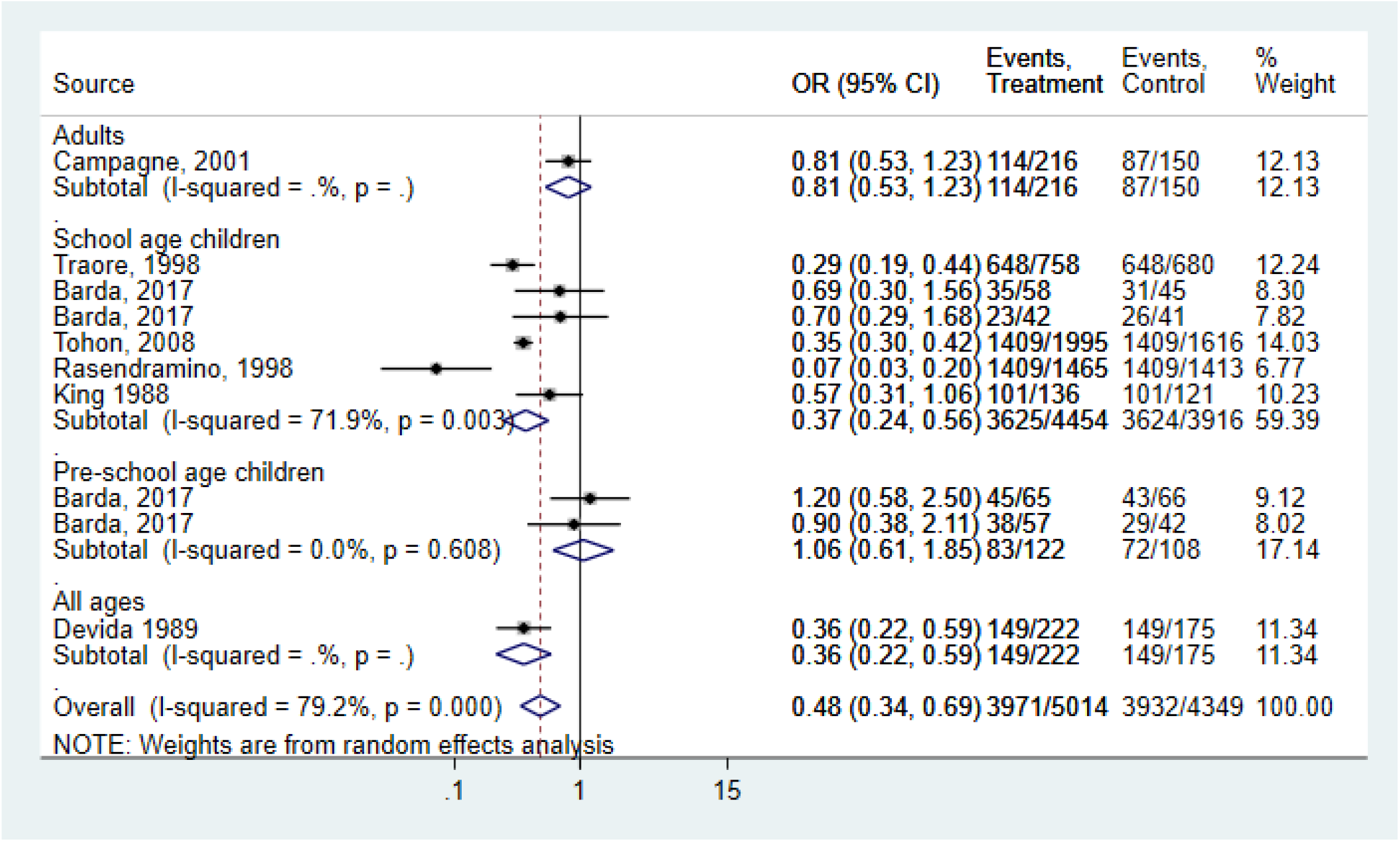
Forest plot for the relation between PZQ/PZQ in combination with other drugs and the risk of urinary tract pathology assessed by population type

#### Urinary bladder lesions

A total of 14 studies provided 15 data for the quantitative analysis of the prevalence of urinary bladder morbidity (Fig. 14). No study reported on PSAC. No significant association of urinary bladder lesions was observed for treatment with PZQ for SAC and adults. But there was a significant reduction in urinary bladder lesions among entire population (0.26, 0.14-0.49; I^2^index=79.1%, p=0.008, n=3). Overall summary OR suggested a 46% (0.54, 0.32-0.90; I^2^index=97.5%, p=0.000, n=15). In individuals treated for *S. haematobium* a reduction in the urinary bladder, lesions were noted (see **Table S3**). No sensitivity analysis was conducted because none of the studies was adjudged moderate or low on RoB for non-randomized controlled trials and with some concerns for randomized controlled trials. The funnel plot was asymmetrically suggesting a potential for a publication bias (see **Fig S1**). This was confirmed in the trim and fill method (0.96, 0.59-1.56; I^2^index=97.3%, p=0.000, Q-statistics=659.51) (see **Table S3**). The certainty of the evidence was low (**Table 1**).

**Fig. 14.**
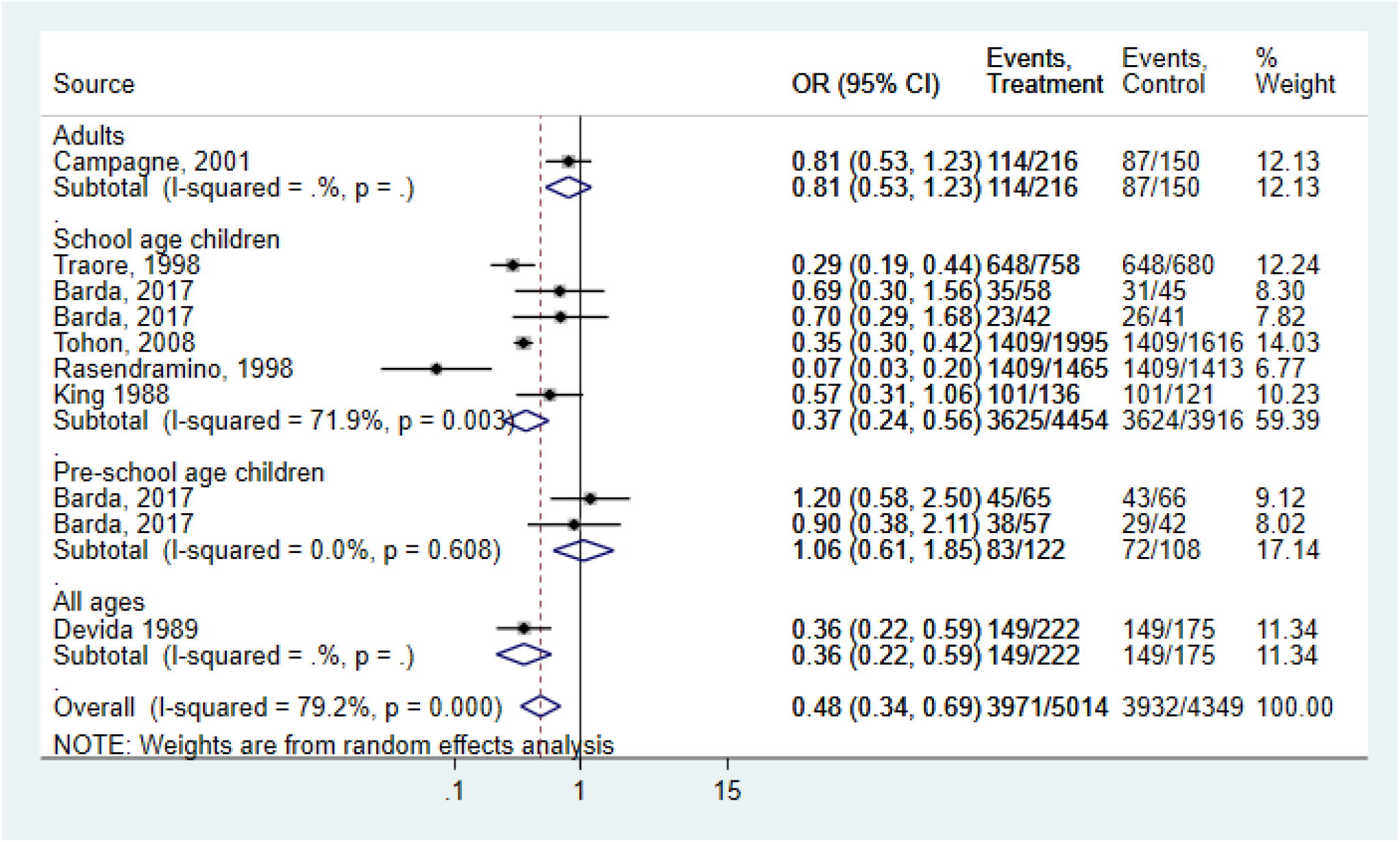
Forest plot for the relation between PZQ/PZQ in combination with other drugs and the risk of Urinary bladder lesions assessed by population type

## Discussion

### Main findings

This systematic review and meta-analysis address the effectiveness of PZQ PC in specific subgroups of the population and can help identify key population group (s) that should be targeted to achieve maximum benefit for the control of infection-related morbidity. A total of 59 studies were identified.

PC treatment with PZQ resulted in reductions in blood in stool in the entire population, haematuria in SAC, proteinuria in PSAC, SAC, and entire population, in portal vein change in SAC, in right lobe hepatomegaly in SAC; in urinary tract pathology in SAC, and in bladder lesion in the entire population. No significant reduction was noted concerning periportal fibrosis, left-sided hepatomegaly, no specific lobe hepatomegaly, and diarrhea prevalence in all age groups. The findings reported here are relevant to many countries in the developing world where individuals continue to be exposed to schistosome species. However, high statistical heterogeneity was observed in some of our subgroup analyses. The certainty of the evidence was very low to low according to GRADE guidelines. Thus, care should be taken in drawing any conclusion that treatment with PZQ for infection-related morbidity is beneficial.

### Methodological validity

The strengths of this systematic review relate to the comprehensive search and collaboration with WHO content experts. There are however several limitations. The majority of the included studies were non-randomized, before-and-after designs without control or repeated cross-sectional designs. Data were unavailable for certain age groups for several key morbidity outcomes (e.g. anemia, hepatomegaly-right-sided lobe, left-sided lobe, and no specific lobe, splenomegaly). The sample size in most analyses was small, leading to important imprecision.

The original studies also applied different interventions such as administration of single doses of PZQ or repeated doses of PZQ in combination with or without other drugs thus, making it difficult to relate PZQ to changes in morbidity outcomes. Differences in responses to treatment may also exist across study populations, and these could be potential sources of the observed heterogeneity. We lacked data on these factors and we also did not have sufficient data from the original studies to undertake meta-regression to ascertain the reasons for heterogeneity.

### Comparison with previous studies

Two recent systematic reviews and meta-analyses have examined the relationship between PC and morbidity outcomes [8, 19]. In the first study, Andrade et al [8] used data from randomized and non-randomized trials for preventive chemotherapy with PZQ or oxaminiquine or metrifonate or hycanthone alone or in combination against soil-transmitted helminthiasis and schistosomiasis. The authors reported large reductions in intermediary morbidity outcomes (e.g. diarrhea: 50%, blood in stool: 79%, protein in urine: 88%, blood in urine: 84%) and small reductions in true morbidity outcomes (e.g. right-sided hepatomegaly: 47%, splenomegaly: 39%, periportal fibrosis: 48%, urinary bladder lesions: 75%) in populations consisting of all age groups. No significant reduction in left-sided hepatomegaly, hepatomegaly with no specific lobe dilated portal vein was observed in this population group. With respect to SAC, large reductions was noted in true morbidity outcomes (e.g. right-sided hepatomegaly: 61%, left-sided hepatomegaly: 63%, no specific lobe hepatomegaly: 87%, splenomegaly: 36%, urinary bladder lesions: 92%) and intermediary morbidity outcomes (e.g. diarrhea: 50%, blood in stool: 79%, protein in urine: 88%, blood in urine: 84%).

The current review identified 46 studies on morbidity outcomes. Unlike Andrade et al [8], we used data from studies focusing on PC treatment with PZQ alone or PZQ in combination with other drugs. Besides, we concentrated on infection-related morbidity outcomes in mostly SAC and the entire population and some data on PSAC and adults with very low or low certainty of the evidence for infection-related morbidity outcomes. We observed large reductions in the prevalence of *S. mansoni* in adults (79%) and SAC (70%) but not in PSAC (39%) and the entire population (46%). Concerning *S. haematobium*, large reductions in infection were observed in PSAC (75%), compared to SAC (67%) and the entire population (68%). We observed large reductions in proteinuria (90%) in PSAC but no data/insufficient data was available for hematuria, diarrhea, and blood in stool in this population. In SAC, significant reductions were noted for hematuria (60%) and proteinuria (64%) but data was lacking for diarrhea and blood in the stool. No significant reduction was observed for the dilated portal vein in the entire population and no data was available for periportal fibrosis no specific lobe hepatomegaly and left-sided hepatomegaly in this population. Concerning dilated portal vein, periportal fibrosis, no specific lobe hepatomegaly, and left-sided hepatomegaly data were lacking in PSAC, SAC, and adults. 57% reduction in right-sided hepatomegaly and 63% reduction in urinary tract pathology was seen in SAC but no significant reduction was noted for splenomegaly and urinary lesion in this population. Data was insufficient/unavailable for PSAC for right-sided hepatomegaly, splenomegaly, urinary tract pathology, and urinary bladder lesion. Like previous studies, Welch et al [19] used data from randomized and non-randomized trials, however, concentrating on mass administration of any drug for chemoprevention of soil-transmitted helminths or schistosomiasis alone or in combination with other deworming drugs. The authors focused on PSAC and SAC. The relevant analysis to the present study was from a sub-group analysis that examined preventive chemotherapy with albendazole/mebendazole alone and in combination with other drugs for schistosomiasis alone in an unspecified age group on outcome of weight, height, and cognition. There was a shred of suggestive evidence that preventive chemotherapy for schistosomiasis may slightly increase weight (0.41 kg, 95% CI −0.20, 1.01; N=1 study) with no change in height with low certainty of evidence. Treatment with PZQ did not affect cognition with moderate certainty of evidence.

There are discrepancies in results between all three meta-analyses. A large part of the discrepancies is due to the inclusion criteria. Whereas the current study focused on PC treatment with PZQ, [8] concentrated on drugs (such as PZQ or oxaminiquine or metrifonate or hycanthone alone or in combination) used in treating soil-transmitted helminthiasis and schistosomiasis. We, however, included all the studies on PZQ in Andrade et al (2017) in our current analysis, and thus, and discrepancies between the current study and that of Andrade et al [8] may due to additional studies on other drugs in their study. It is also important to note that Welch et al [19] also included only one study on treatment with PZQ. The infection morbidity reduction found in the meta-analyses was largely based on observational studies where residual confounding and selection bias were serious problems. Where conducted randomized control trials produced null or marginal reduction.

Welch et al [19] targeted PC treatment in SAC and Andrade et al [8] focus on PC treatment in SAC and the entire population. We targeted PC treatment in PSAC, SAC, adults, and the entire population. Identifying the ideal age for PC treatment against schistosomiasis is quite challenging. Expanding PC programs across all age groups has a direct benefit to the targeted population also indirectly reduces overall transmission across all age groups [20, 21]. The latter is not usually captured and was not considered in the systematic reviews. Many of the study outcomes in the systematic reviews do not directly measure morbidity but rather are intermediate measures of pathology. Again, the relationship between preventive chemotherapy and disease outcomes remains complex given the mix of acute reversible and chronic irreversible pathology, different Schistosoma species, and the consideration of epidemiologic complexities of transmission that may limit the ability to detect generalized findings. Scarcity of data coupled with lack of high-quality data limit on this topic, limit any clinical meaningful relationship in key age groups. Evidence to support PC in SAC based on parasitological and some morbidity outcomes are consistent in all the systematic reviews but are of low quality. The evidence to support consideration of other key age groups in PC programs to reduce parasitological and morbidity outcomes is also of low quality.

## Conclusion

In conclusion, PC with PZQ reduces the prevalence of *S. mansoni* and *S. haematobium* infections and reduces some morbidity outcomes in SAC. There is suggestive evidence that PSAC and adult populations may benefit from some outcomes. This assertion was based on low certainty of evidence.

## Data Availability

All data produced in the present study are available upon reasonable request to the authors

## Abbreviations

AHRQ: Agency for Healthcare Research and Quality
CI: Confidence Interval
MDA: Mass Drug Administration
OR: Odds Ratio
PC: Preventive Chemotherapy
PICO: population, intervention, comparison, and outcome..
PRISMA: Preferred Reporting Items for Systematic Reviews and Meta-Analyses statement
PSAC: Pre-School Age Children
PZQ: Praziquantel
RCTs: Randomized Control Trials
RoB-I/GRADE: Grading of Recommendations, Assessment, Development, and Evaluation ….
SAC: School-age children
WHA: World Health Assembly
WHO: World Health Organization

## Ethics approval and consent to participate

Not applicable

## Consent for publication

Not applicable

## Availability of data and materials

Available on request from RQ

## Authors’ contributions

RQ & TDA developed the search strategy. RQ screened all articles, supervised the work, and drafted the first version of the manuscript. AY, ABC, BO, and EA extracted the data. RQ, TDA, HMM, AGD, and NL conceptualized the research questions and interpreted the result of the final version of the manuscript. All authors read and approved the manuscript.

## Acknowledgment

We thank all authors of the eligible studies

